# Translating *SUVR* bias correction to amyloid PET enables early imaging and more accurate simplified quantification

**DOI:** 10.64898/2026.08.24.26361268

**Authors:** Praveen Honhar, Michael Properzi, Aaron Schultz, Keith A Johnson, Julie C Price

## Abstract

**Introduction:** A new method that corrects for time-dependent bias in standardized-uptake value ratios (SUVRs) was adapted and optimized for [^11^C]PiB (PiB) amyloid-beta (Aβ) PET, across low-to-high Aβ loads, relying only on PET data collected during the *SUVR* time-window. This modeling approach was evaluated in cross-sectional and longitudinal cohorts for earlier and shorter SUVR time-windows (30-45 min, 45-60 min) than commonly applied, to enable higher throughput imaging.

**Methods:** The *SUVR* correction (*SUVR*c) approach was optimized and tested on separate cross-sectional (n=88), and longitudinal (36 participants, two time-points, 72 images) cohorts from the Harvard Aging Brain Study. The cross-sectional cohort spanned low, intermediate and high levels of cortical Aβ pathology and the longitudinal images included two cohorts with low (5-10%) and high levels (~40%) of Aβ change. *SUVR* and *SUVR*c were compared against SRTM *DVR* (0-60 min) to quantify Aβ burden through Pearson’s and Lin’s correlations, difference plots and longitudinal change.

**Results:** The mean regional bias in PiB *SUVR* (5-15%, depending on time-window and Aβ burden) was significantly reduced to < 3% by *SUVR*c (corrected *p* < 0.05) in the cross-sectional cohorts for all time-windows, along with reductions in bias variability. *SUVR*c also showed higher Pearson’s correlation (r) and Lin’s concordance (LCC) with *DVR* across time-windows (r=0.98, LCC=0.99 at 30-45 min and 45-60 min) compared to uncorrected *SUVR* (r=0.96, LCC=0.95 at 30-45 min, r=0.97, LCC=0.92 at 45-60 min). Bland-Altman plots confirmed better agreement between *SUVR*c and *DVR* (mean bias at 30-45 min: 0.02 for *SUVR*c, 0.10 for *SUVR*; mean bias at 45-60 min: 0.01 for *SUVR*c, 0.17 for *SUVR*). Longitudinal *DVR* changes were more accurately represented by *SUVR*c, compared to uncorrected *SUVR*.

**Conclusions:** *SUVR*c for [^11^C]PiB PET enables more accurate quantification of Aβ burden than *SUVR* in cross-sectional and longitudinal studies (relative to SRTM DVR), while enabling imaging at earlier and shorter time-windows. The improved accuracy would be beneficial in better quantifying amyloid re-emergence post anti-amyloid therapy and could be used for kinetic harmonization across time-windows and radiotracers.

## Introduction

Positron emission tomography (PET) imaging of amyloid-β (Aβ) deposition, enabled by radiotracers such as [^11^C]Pittsburgh Compound-B ([^11^C]PiB), can play a central or secondary role in the diagnosis of Alzheimer’s disease (AD)^1^, as well as in recruitment and endpoint design of AD prevention trials for anti-amyloid therapies^2-5^. Quantitative PET measures are critical for identifying early pathological changes and evaluating therapeutic interventions targeting amyloid accumulation. Distribution volume ratio (*DVR*)^6^, derived from the simplified reference tissue model (SRTM)^7^ or Logan reference graphical method^8^, is generally regarded as the gold-standard quantitative index of amyloid burden^9-11^ (in absence of arterial sampling) because it accounts for tracer delivery and non-equilibrium radiotracer clearance, but its estimation requires ~ 60 min of dynamic imaging data and kinetic modeling, which is impractical for routine clinical or large-scale trial use. *SUVR* computed as the ratio of average tracer uptake in a target region to a reference region during a predefined late-imaging time-window, is therefore a widely utilized PET outcome.

Numerous studies have demonstrated that *SUVR* measurements can be biased relative to *DVR* due to regional differences in radiotracer washout rates^12,13^. For [^11^C]PiB, this bias will be positive and higher for later *SUVR* time-windows, due to its fast kinetics. Previous [^11^C]PiB studies have reported 40-60 min and 50-70 min as optimal *SUVR* choices^14^, based on correlation with *DVR*. However, (1) *SUVR* bias is a function of radiotracer clearance, resulting in a time-window dependent and Aβ load (negligible vs high Aβ) dependent bias; and (2) there is also a remarkable regional-variation in Aβ burden, which influences regional *SUVR* bias, within subject.

Few approaches have addressed these limitations. Two prominent methods include a simplified graphical method called the relative-equilibrium plot^15^ that allows *SUVR* bias correction^16^, and a correction based on kinetic modeling principles known as the tissue clearance correction^17,18^. These methods are applicable only under ‘transient or relative equilibrium’ paradigm; a state where the radiotracer clearance has attained a terminal value that is constant in plasma and brain regions. This assumption holds only at late post-injection times for [^11^C]PiB, and for the graphical relative-equilibrium method, requires late-time dynamic data (typically 55-90 min p.i.^16^). A recent innovation is the development of a generalized version of tissue clearance correction, called *SUVR* correction^19^, that does not require transient equilibrium and can be applied when the radiotracer has different clearance rates in plasma, target and reference regions. This can correct time-dependent *SUVR* bias and can enable imaging at earlier time-windows (higher-throughput amyloid imaging with higher counts for [^11^C]PiB).

Per this strategy, the *SUVR* bias can be corrected as follows^19^,

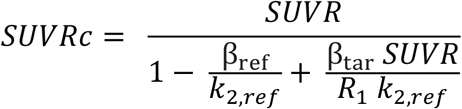

*SUVR*c is the bias-corrected *SUVR*; the denominator represents correction factor for *SUVR* bias due to non-equilibrium radiotracer clearance. βref and βtar are mono-exponential clearance rates of the radiotracer in the reference and target regions during *SUVR* time-window. Note that if βref = βtar =0 (system at equilibrium), no correction is needed. Further, this correction requires *a priori* estimates of *k*2,ref (*k*2 parameter of the reference region) and *R*1 (ratio of radiotracer delivery in target and reference tissues) which are typically supplied as population-averaged values in target-tissue type^19^.

For successful implementation, the estimation of βtar is a crucial factor. Although regional clearance rates can theoretically be estimated directly from dynamic time-activity curves (TACs) acquired during the *SUVR* time window, TAC noise and the high sensitivity of the correction to errors in βtar limit this approach^19^. Previous neuroreceptor PET studies showed that regional clearance rates follow predictable relationships with tracer uptake, enabling empirical models to estimate βtar^19,20^. However, pathological protein targets such as Aβ exhibit a broader range of uptake than conventional neuroreceptor targets, spanning negligible, intermediate, and high cortical deposition across healthy aging, preclinical AD, mild cognitive impairment (MCI), and AD^21,22^. Consequently, refined empirical models are required to estimate βtar across the full continuum of Aβ pathology.

Here, we adapt the *SUVR* correction method to [^11^C]PiB imaging. Bias was corrected within two time-windows between 30-60 min post-injection. The method was optimized across the spectrum of pathology–from low to high Aβ loads enabling generalization. Different empirical models, depending on the global Aβ load, were developed to better estimate βtar. The method was optimized and tested in separate training and validation cross-sectional cohorts from the Harvard Aging Brain Study^23^ (HABS). Performance of both *SUVR* and *SUVR*c were compared against SRTM *DVR* to understand regional bias and variability. Lastly, *SUVR* correction was also tested on two longitudinal cohorts (either low (5-10%) or high (~ 40%) levels of Aβ change) to characterize change over time.

### Study Participants

Imaging data from a subset of participants enrolled in HABS were reanalyzed. A cross-sectional cohort included 88 older adults along a spectrum of Aβ pathology, from negligible to high global burden. Two longitudinal cohorts (n=18/cohort) with two time-points per cohort were also included; one with smaller changes in neocortical Aβ (5-10% increase) and the second with larger changes in Aβ (30-70%). See Table 1 for clinical and demographic characteristics. We divided data by neocortical (NEO) *SUVR* into empirical groups of low (< 1.10), intermediate (1.10 – 1.75), and high (> 1.75) amyloid. This stratification allowed us to develop different empirical models for βtar, depending on amyloid burden.

**Table 1.** Participant Characteristics. Demographic data are presented as mean± SD. Cross-sectional participants were stratified by baseline amyloid burden. Longitudinal participants were stratified by annual longitudinal percent change (LPC).

| Characteristic | Cross-Sectional Cohorts |  |  | Longitudinal Cohorts |  |  |  |
| --- | --- | --- | --- | --- | --- | --- | --- |
|  | Low Amyloid | Intermediate Amyloid | High Amyloid | Low LPC |  | High LPC |  |
|  | NEO SUVR < 1.10 | 1.10 < NEO SUVR < 1.75 | NEO SUVR > 1.75 | Time Point 1 | Time Point 2 | Time Point 1 | Time Point 2 |
| N | 30 | 29 | 29 | 18 | 18 | 18 | 18 |
| Age (Years) | 73±7 | 76±8 | 81±6 | 72±6 | 82±7 | 72±5 | 78±5 |
| Female Sex (N, %) | 24, 80% | 21, 72.4% | 24, 82.7% | 11, 61.1% | - | 12, 66.7% | - |
| Education (Years) | 15.2±3.2 | 16.5±2.6 | 17.2±2.4 | 17.1±2.6 | - | 15.7±3.4 | - |
| PACC5 (Z-Scores) | 0.22±0.87 | 0.02±0.83 | -0.74±1.11 | 0.35±0.52 | -0.41±1.01 | 0.06±0.76 | -0.03±0.77 |
| Clinical Diagnosis | 30 CN | 29 CN | 22 CN, 3 MCI, 4 AD | 18 CN | 16 CN, 2 AD | 18 CN | 17 CN, 1 MCI |

Detailed inclusion and exclusion criteria for the HABS cohort have been previously published^23^. The study procedures were approved by the Institutional Review Board for Massachusetts General Brigham. All participants provided informed consent.

## Materials and Methods

### PET and MR Imaging

Dynamic PET images (0-60 min post injection) were acquired for all participants on a Siemens ECAT HR+ tomograph at Massachusetts General Hospital after a bolus injection of [^11^C]PiB (dose:534±57 MBq). Participants also underwent 3T T1-weighted structural MR imaging at the MGH-Martinos Center for Biomedical Imaging.

### PET Processing and Kinetic Modeling

PET data were reconstructed using an ordered-subset expectation-maximization (OSEM) algorithm (8x15s,4x60s,27x120s) with corrections for scatter, randoms, attenuation and radioactive decay. Frame-to-frame motion correction was performed using the Statistical Parametric Mapping’s (SPM12) Realign tool. Freesurfer (v6.0) was used to segment T1 images into cortical (aparc) regions using the Desikan-Killiany atlas^24^. Eleven regions-of-interest (ROIs) defined within the cerebral cortex, including a previously described neocortical composite were analyzed^25,26^. Based on past studies, cerebellar gray matter was used as the reference region^27^. ROIs were transformed to the native PET space to extract time activity curves (TACs) for the target and reference regions. *SUVR* values were computed for target ROIs over 30-45 min and 45-60 min time-windows and regional *DVR* values for all regions were computed as *BP*ND + 1 from SRTM fits to 60-min dynamic data (single-step SRTM used for high Aβ images, a version of SRTM2 used for intermediate and low Aβ images, see supplementary material for implementation details).

### Empirical Models for Estimating [^11^C]PiB Clearance Rates during *SUVR* Time-Windows

For the reference region-cerebellar cortex (a region with large volume and low noise), βref was directly estimated as the rate constant from a mono-exponential fit to the reference region TAC during the *SUVR* time-window (8 time-points). Theoretically βtar can also be estimated in this manner, however, noise in target TACs (especially in smaller ROIs or voxel level) limit this technique. Previous neuroreceptor studies^19,20^ addressed this using the following empirical model for estimation of βtar,

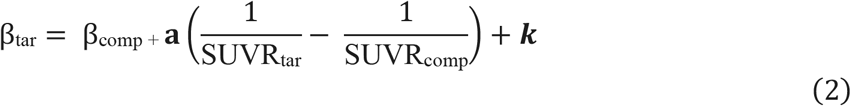

This model assumes the existence of at least one composite region (comp) with a large volume (low noise) and high radiotracer uptake whose clearance rate (βcomp) can be directly estimated from its TAC during the *SUVR* time-window. The model underscores the inverse relationship between βtar of a region and its radiotracer uptake in *SUVR* units, i.e., regions with higher uptake than the composite region will have a slower clearance rate and vice-versa. **a** and ***k*** are parameters that must be estimated from a pilot cohort and depend on the radiotracer as well as the time-window. Previous neuroreceptor PET studies fixed *k*=0 so that βtar = βcomp when the target and composite regions are identical^19,20^. However, unlike neuroreceptor PET, several cortical subregions in amyloid PET may exhibit higher uptake than the neocortical composite region. Therefore, estimating *k* from the training data better optimizes prediction of βtar over multiple regions.

We used the model described in equation-2 for estimating βtar in the ‘high amyloid’ cohort (NEO *SUVR* > 1.75, n=29). First, we randomly divided the high amyloid cohort into a training (n=9) and a validation (n=20) set. For the training set, regional βtar values were estimated using SRTM fits to regional TACs. The training set was then used to estimate **a** and ***k*** in equation 2 using a least-squares minimization approach, with separate values for these parameters estimated for the 30-45 min and 45-60 min time-windows.

A crucial difference between neuroreceptor and pathological protein studies is the much wider spectrum of pathological burden in the human brain. For example, older adults with normal cognition or preclinical Alzheimer’s disease may have negligible to small levels of Aβ, for which the composite region-based paradigm would be sub-optimal. Therefore, alternate models were needed to sufficiently describe βtar in participants with different levels of Aβ pathology. We propose the following models for ‘low amyloid’ and ‘intermediate amyloid’ groups.

### Low Amyloid Group (NEO *SUVR* < 1.10)

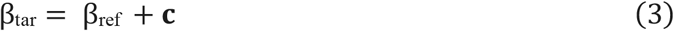

### Intermediate Amyloid Group (1.10 < NEO *SUVR* <1.75)

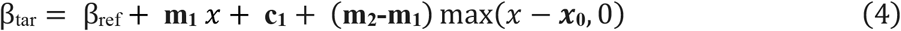

where,

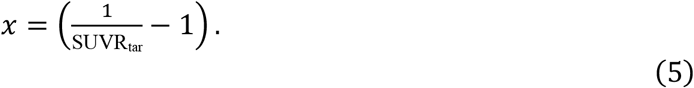

For the low amyloid group, the model approximates βtar as βref plus a constant term. The rationale behind this choice is that in the absence of or low levels of amyloid most TACs in gray matter regions should be similar to the TAC of the reference region. Next, for the intermediate amyloid levels, βtar is modeled as βref plus a continuous two-segment piecewise linear function of *x* = (1/SUVRtar − 1), with slopes **m**_**1**_ and **m**_**2**_, knot location ***x***_**0**_, and first-segment intercept **c**_**1**_. This model is similar to equation 2, but with the deviations in 1/*SUVR*tar computed relative to reference region rather than a composite region, and different values of the radiotracer constants (**m**_**1**_ and **m**_**2**_) depending on how high (or low) *SUVR*tar is instead of using a single value (**a**) throughout. This is consistent with the heterogeneity of the intermediate group, with *SUVR*tar at the lower end of the group being similar to the low amyloid group and higher values deviating significantly from it. It should be noted that for a target region with *SUVR*tar values close to 1 (low amyloid), equation 4 reduces to equation 3 as terms with (1/*SUVR*tar − 1) and ***x***_**0**_ drop out.

The parameters **c** and **[m**_**1**_, **m**_**2**_, ***x***_**0**_, **c**_**1**_] must also be estimated from the low and intermediate cohort pilot data, respectively, similar to how **a** and ***k*** were estimated in the high amyloid group. We followed the same setup of dividing their respective cohort data into a training and validation set (1:2 ratio), with parameters estimated through a least-squares minimization algorithm on training data, separately for 30-45 min and 45-60 min time-windows.

### Comparison of *SUVR, SUVR*c and *DVR* in the Cross-Sectional Validation Cohort

Regional *SUVR* values for all target ROIs were corrected using equation 1 in the three cross-sectional validation cohorts (low, intermediate and high amyloid) to yield corresponding *SUVR*c values. For the correction, βtar during a given *SUVR* time-window was estimated from the empirical models described in equations 2-4, while βref was estimated through a mono-exponential fit to the reference region TAC. Fixed population-averaged values of *k*2’= 0.054±0.009 min^−1^ (mean across individuals) and *R*1 = 0.93±0.091 (mean across individuals and ROIs) were used in the correction formula.

Percent bias in *SUVR* and *SUVR*c were characterized as a percentage of *DVR* as:

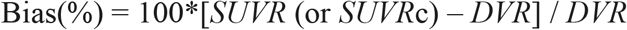

Mean percent bias in *SUVR* and *SUVR*c and SD of percent bias were compared regionally, across the three cross-sectional cohorts (validation datasets) and two-different time-windows.

Results were then pooled across all three cohorts and Bland-Altman difference plots were used to characterize the difference between *SUVR* and *SUVR*c across the entire continuum of amyloid burden. Finally, Pearson’s correlations and Lin’s concordance coefficients were also characterized between *SUVR* and *DVR*, and *SUVR*c and *DVR*.

### Comparison of *SUVR, SUVR*c and *DVR* for Measuring Longitudinal Change

Longitudinal percent change (LPC) over two time-points was defined as:

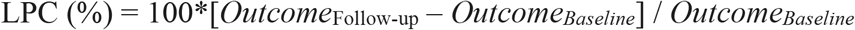

where ‘Outcome’ could be *DVR, SUVR* or *SUVR*c. Mean regional LPC (across participants) were compared across all outcomes, in two separate longitudinal cohorts, henceforth, small-LPC (n=18, 5-10% Aβ increase) and large-LPC (n=18, 30-70% Aβ increase) cohorts. Each scan was characterized as low, intermediate or high amyloid depending on the NEO *SUVR* criteria discussed earlier and this categorization was used to select the empirical model for βtar, consistent with cross-sectional analyses.

## Results

### Empirical Models for Estimating [^11^C]PiB Clearance Rates during *SUVR* Time-Windows

Model parameters were reliably estimated from training datasets for low, intermediate and high amyloid groups across two time-intervals (30-45 min and 45-60 min). These values are presented in Table 2. All estimated parameters had < 40% standard error (s.e.), demonstrating robust identification, except for the second slope (m2) in the two-segment piecewise linear model for β_tar_ in the intermediate amyloid group, where the large s.e. resulted from m2 being close to zero compared to the slope of the first segment (m1).

**Table 2.**
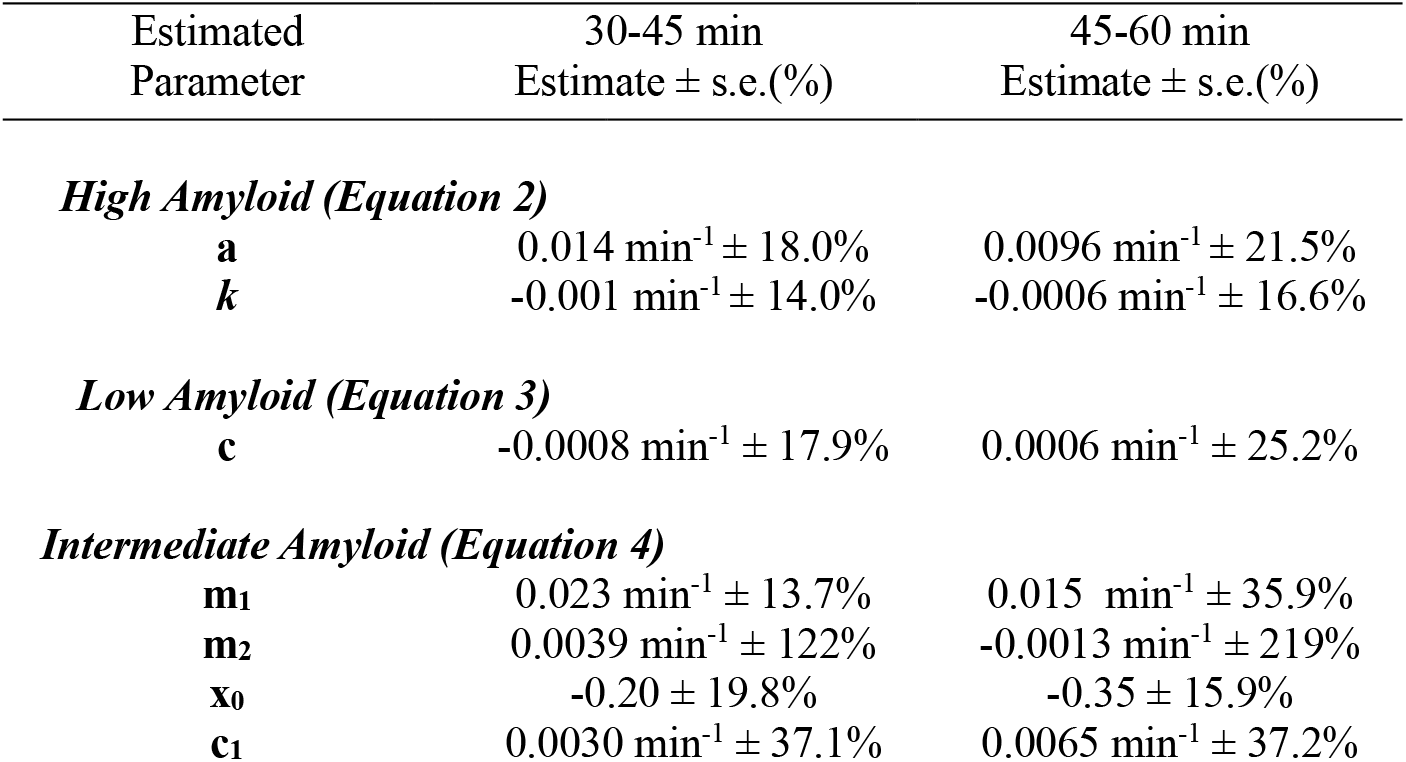
Estimated values of model parameters ± standard error (%) for β_tar_ prediction in training data.

### Comparison of *SUVR, SUVR*c and *DVR* in the Cross-Sectional Validation Cohort

On average, across subjects and ROIs, *SUVR* was positively biased in all three cross-sectional cohorts at both time-points compared to *DVR* (5.6±6.3%, 8.0±2.6%, 4.8±3.8% at 30-45 min and 14.7±5.8%, 13.0±4.3%, 6.9±4.2% at 45-60 min for the high, intermediate and low amyloid cohorts, respectively). *SUVR*c had lower bias and bias variability, for both time-points (on average: 1.6±5.2%, 0.87±2.6%, 0.64±2.5% at 30-45 min, and −0.15±4.5%, 2.2±2.5%, 2.2±3.5% at 45-60 min, respectively, for the high, intermediate and low amyloid cohorts). Figures 1 and 2 compare mean regional bias and its variability (in units of percent *DVR*) for *SUVR* and *SUVR*c at the two time-windows respectively. The reduction in bias in the high amyloid cohort was statistically significant (paired *t*-tests) for 8 /11 regions at the 30-45 min time-window (corrected-pFDR < 0.05), and for all regions (corrected-pFDR < 1×10^−9^) for 45-60 min. The bias reduction was also statistically significant for both time-points for the intermediate and low amyloid groups (corrected-pFDR < 0.01).

**Figure 1.**
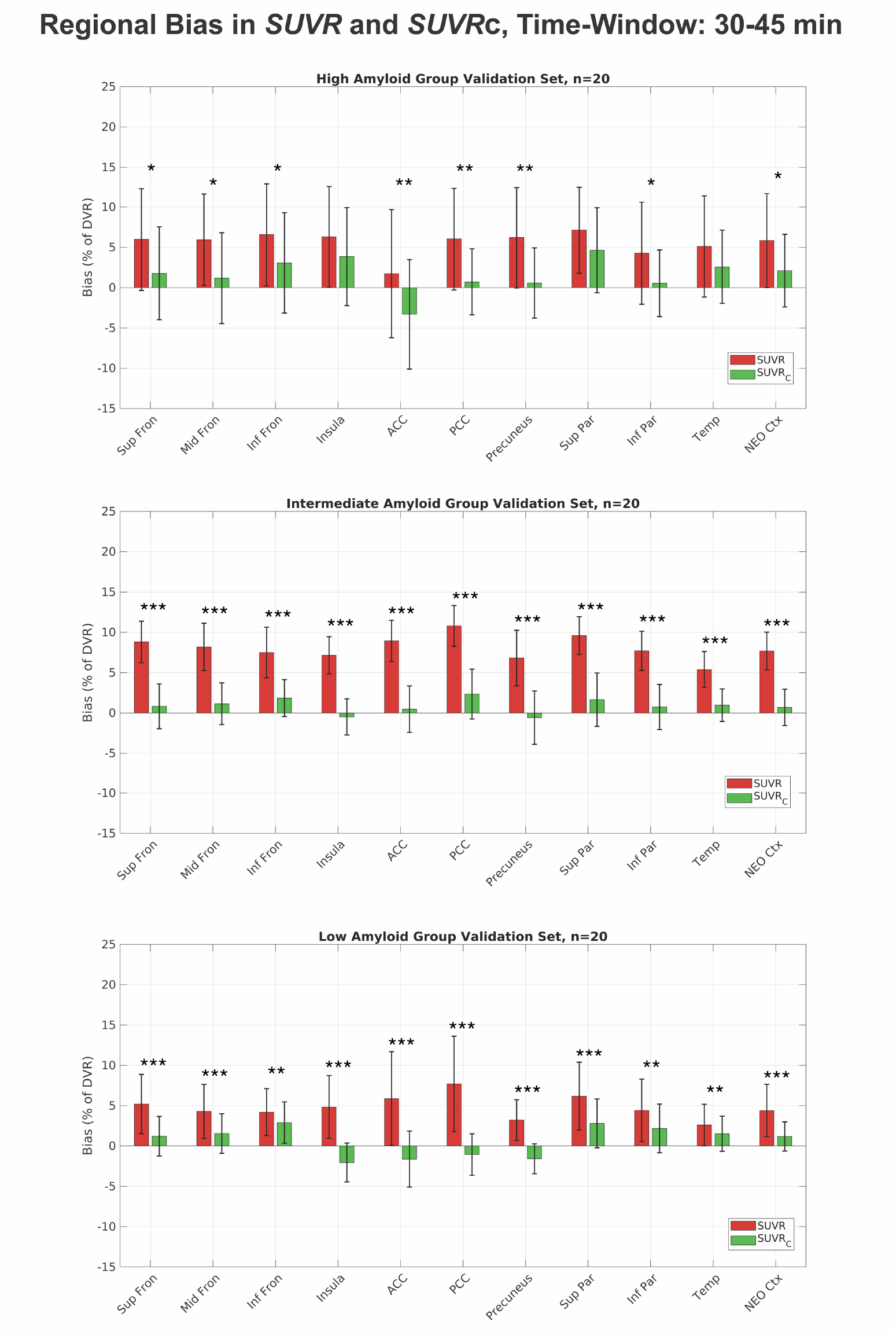
*SUVR* correction significantly reduced regional [^11^C]PiB *SUVR* bias and its SD at 30-45 min across the spectrum of amyloid burden. * corr-p_FDR_<0.05, ** corr-p_FDR_<0.01, *** corr-p_FDR_<0.001.

**Figure 2.**
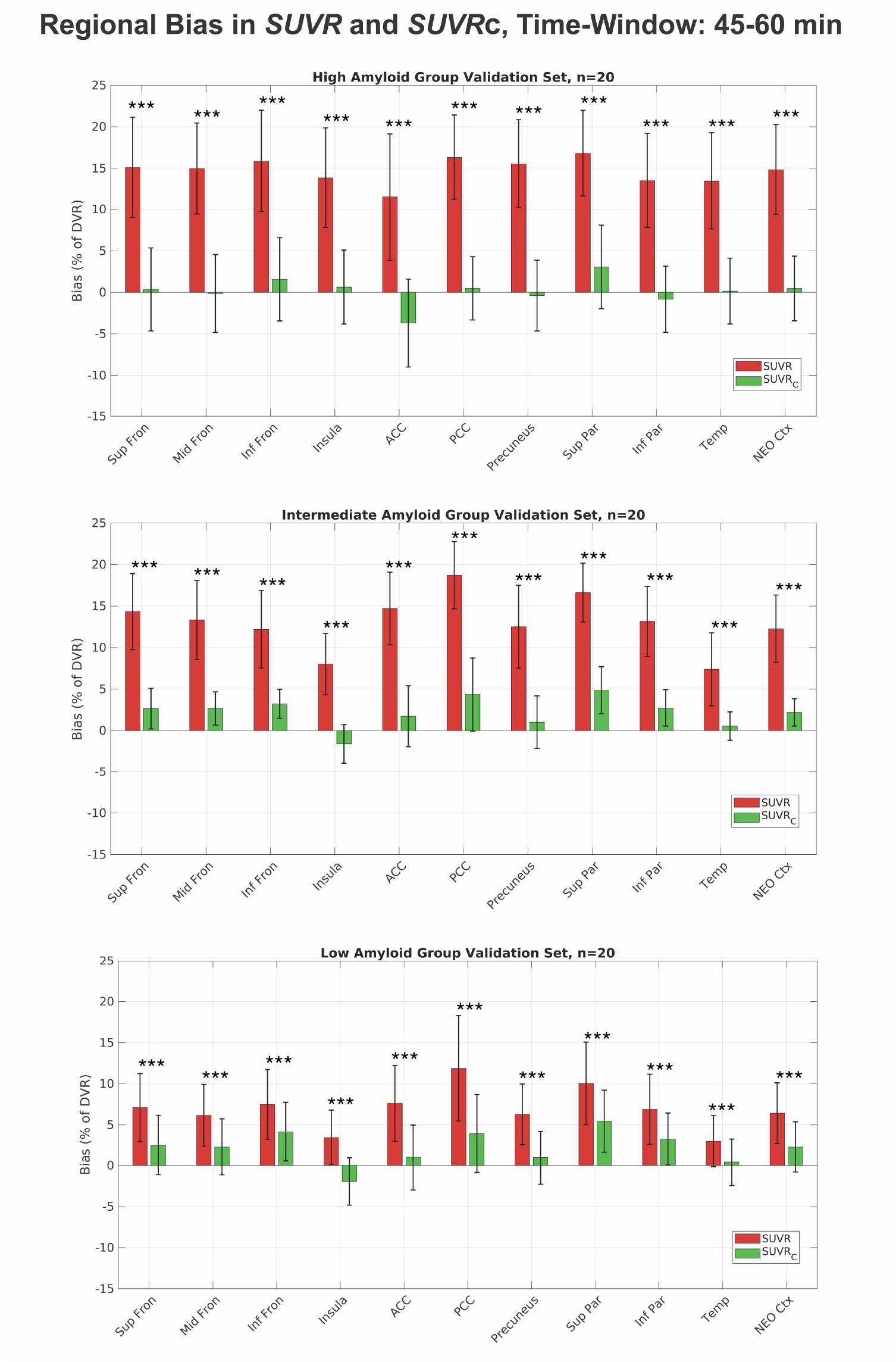
*SUVR* correction significantly reduced regional [^11^C]PiB *SUVR* bias and its SD at 45-60 min across the spectrum of amyloid burden. *** corr-p_FDR_<0.001.

Bland-Altman analyses comparing *SUVR* and *SUVR*c with *DVR* across participants and ROIs, showed a smaller difference between *SUVR*c and *DVR* (mean difference: 0.018 at 30-45 min and 0.014 at 45-60 min, with corresponding widths for limits of agreement [2 x 1.96 x SD of differences]: 0.28 at 30-45 min and 0.26 at 45-60 min, Figure 3). Higher differences were observed between *SUVR* and *DVR* (mean bias: 0.104 at 30-45 min and 0.174 at 45-60 min, with higher widths for limits of agreement: 0.40 at 30-45 min and 0.51 at 45-60 min, Figure 3). These trends were also observed at a single-ROI level (Supplementary Figure 1).

**Figure 3.**
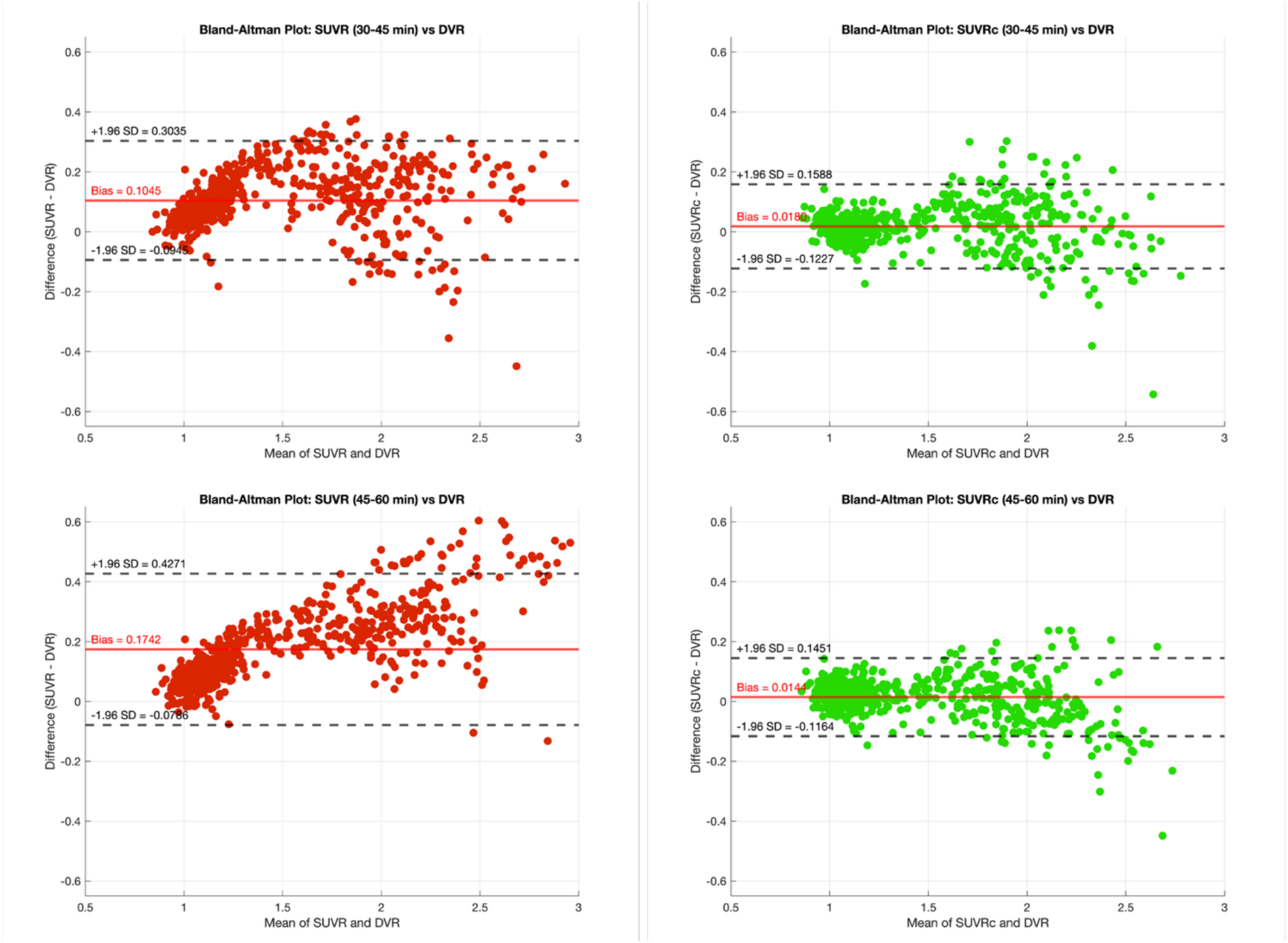
Bland-Altman analyses of *SUVR* (left) and *SUVR*c (right) with *DVR* across all cross-sectional participants (n=88) and ROIs (m=11) show better agreement between *SUVR*c and *DVR*. Additionally, *SUVR* bias (y-axis) values is a function of *DVR* itself, but this confound is also absent in *SUVR*c.

Pearson’s correlation coefficient between *DVR* and *SUVR*c across subjects and ROIs was slightly higher than *SUVR* (Pearson’s R^2^: 0.956 for *SUVR* vs 0.976 for *SUVR*c at 30-45 min and 0.975 for *SUVR* vs 0.981 for *SUVR*c at 45-60 min, Figure 4). Lin’s concordance correlation coefficient between *DVR* and *SUVR*c was higher than between *DVR* and *SUVR* (Lin CCC: 0.954 for *SUVR* vs 0.987 at 30-45 min and 0.918 for *SUVR* vs 0.989 for *SUVRc* at 45-60 min), demonstrating superior agreement between *DVR* and *SUVR*c. These trends were also observed at a single-ROI level (Supplementary Figure 2).

**Figure 4.**
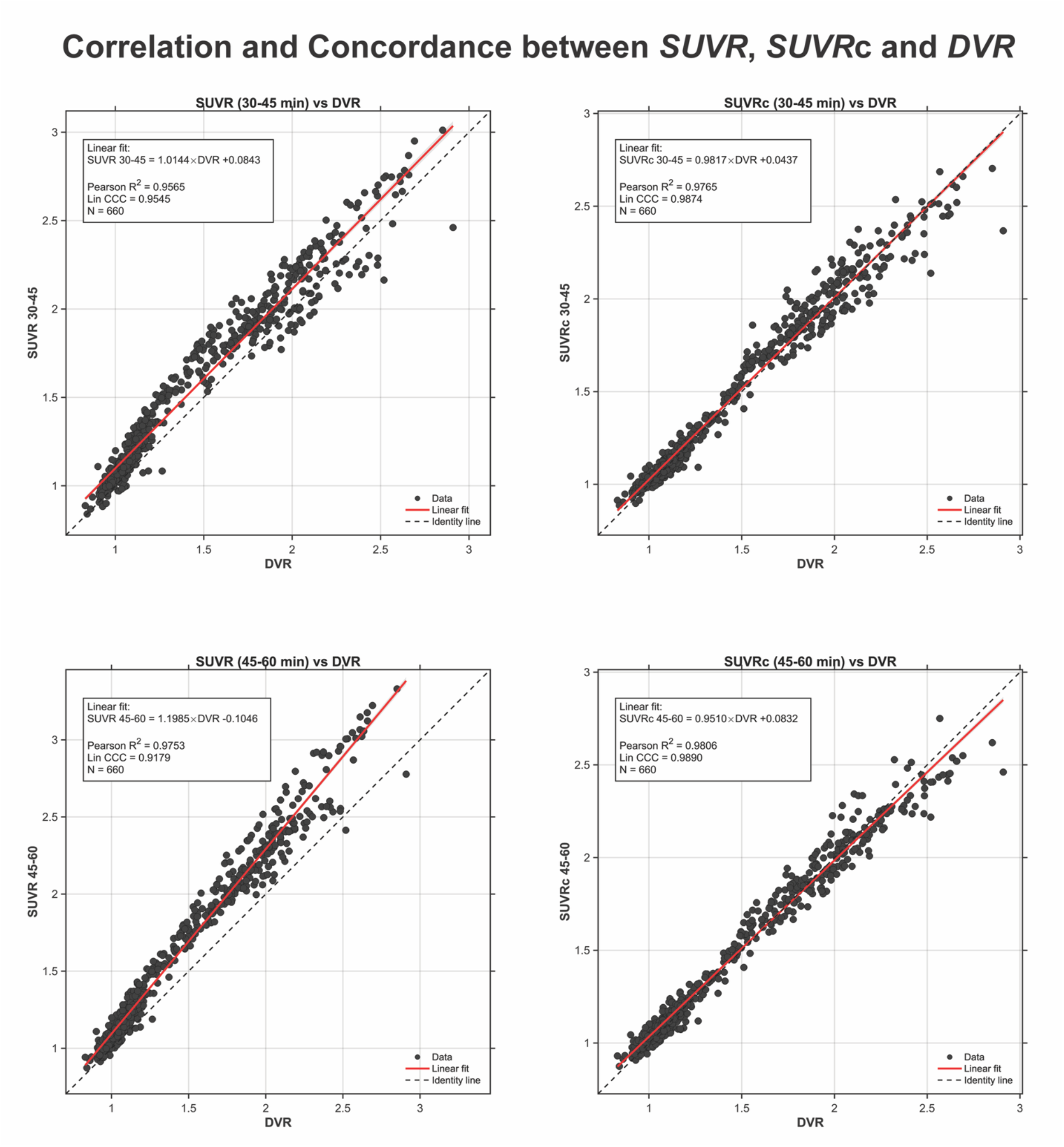
Pearson’s correlation and Lin’s concordance coefficients of *SUVR* (left) and *SUVR*c (right) with *DVR* across all cross-sectional participants (n=88) and ROIs (m=11) show better correlation and improved concordance between *SUVR*c and *DVR* that does not depend on the time-window.

### Comparison of *SUVR, SUVR*c and *DVR* for Measuring Longitudinal Change

In the Small-LPC cohort (5-10% longitudinal change), the best correspondence between a simplified PET outcome and *DVR* change was observed for *SUVR*c at 30-45 min time-window (Mean LPC for neocortex ± s.e.m : 6.2±0.9% for *DVR*, 7.9±0.8% for *SUVR* and 6.3±0.9% for *SUVR*c). *SUVR* consistently over-estimated amyloid deposition in this cohort, while regional-LPC derived from *SUVR*c was more consistent with *DVR* (Figure 5). In the Large-LPC cohort (30-70% longitudinal change), *SUVR*30-45min based-LPC underestimated *DVR-*based LPC for all regions, while *SUVR*c30-45min-based LPC was more similar to *DVR* LPC (Figure 5, Mean LPC for neocortex ± s.e.m : 43.2±3.4% for *DVR*, 34.4±3.3% for *SUVR* and 40.0±3.7% for *SUVR*c). At 45-60 min time-window, both *SUVR* and *SUVR*c-based LPC considerably overestimated amyloid change in the Small-LPC cohort, but the bias was larger for *SUVR* (Mean LPC for neocortex±s.e.m : 6.2±0.9% for *DVR*, 9.7±0.9% for *SUVR* and 8.3±1.0% for *SUVR*c). Finally, both *SUVR* and *SUVR*c at 45-60 min showed comparable performance in measuring large amyloid changes, with either metric slightly underestimating *DVR*-based changes (Supplementary Figure 3). Taken together, these results imply a more accurate quantification of Aβ change by *SUVR*c, compared to raw *SUVR* values.

**Figure 5.**
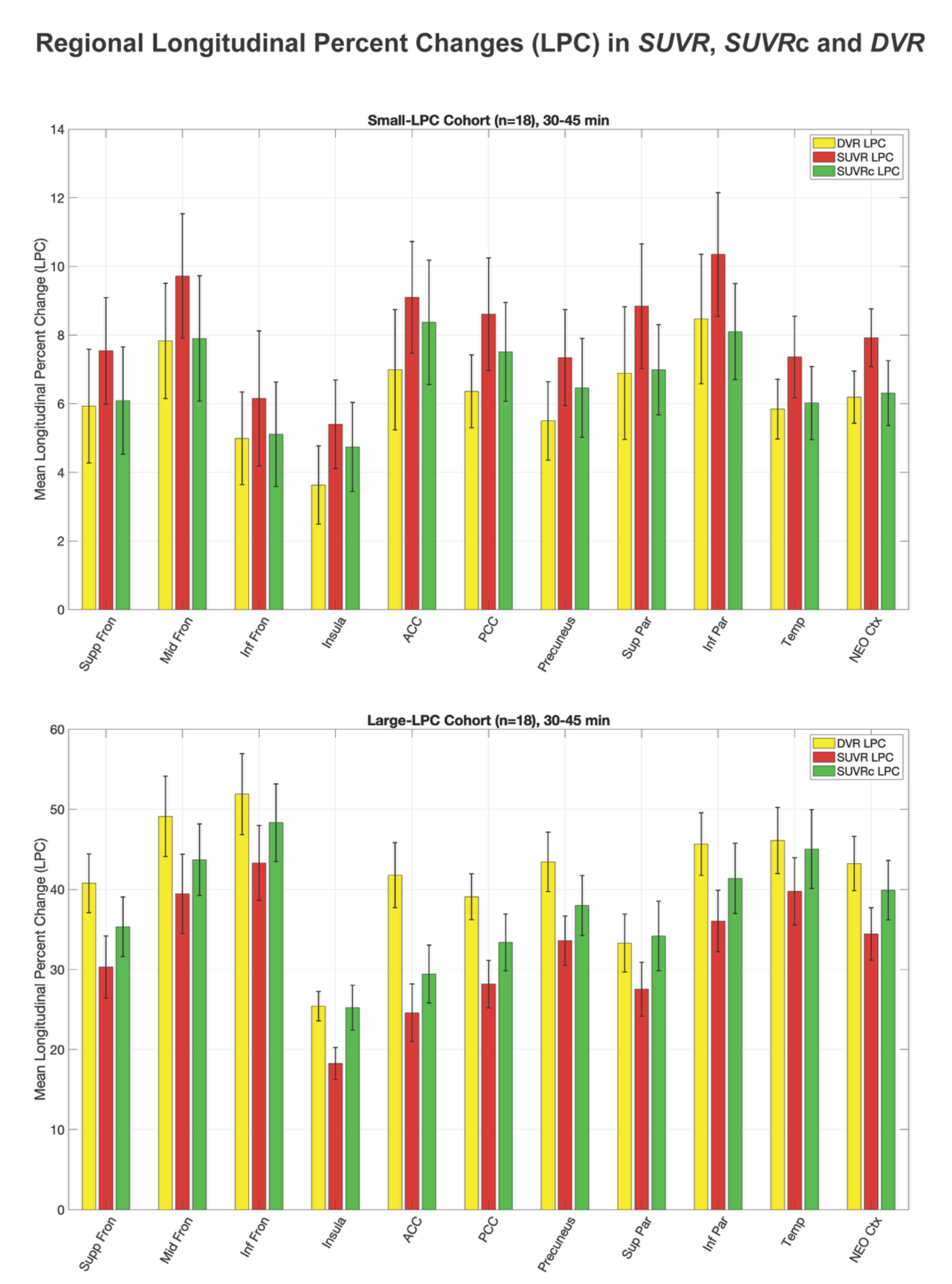
Mean longitudinal percent changes in *SUVR*c 30-45 min showed the best concordance with true changes (based on *DVR*) for the small and large longitudinal cohorts. *SUVR*c improved results compared to *SUVR*. Data shown are mean±s.e.m.

## Discussion

We adapted and successfully optimized the *SUVR* correction approach for amyloid PET using [^11^C]PiB as a more accurate and precise measure of Aβ burden than *SUVR*. The correction paradigm is designed to operate during non-equilibrium conditions after a bolus injection, when regional radiotracer clearance rates vary across regions. The performance of the correction was optimized and tested in independent samples across the full spectrum of Aβ pathology – from negligible to high levels – and *SUVR*c demonstrated a lower bias and often a lower variance in regional bias, compared to ground-truth *DVR*. Relying only on PET data collected during the *SUVR* time-window, this method dramatically improves the concordance between *SUVR* and *DVR* at a single-scan level.

The correction of *SUVR* bias at 30-45 min time-window shortens amyloid PET protocols and enables higher-throughput imaging in hospital settings. For C-11 based radiotracers, scanning early also improves image statistics via higher count rates. Similar performance of *SUVR*c in correcting *SUVR* bias during 30-45 min and 45-60 min indicate utility of the correction for cross-center harmonization from studies employing different time-windows.

Beyond bias reduction in cross-sectional studies, [^11^C]PiB *SUVR*c enabled more accurate measurement of longitudinal change in Aβ burden. Raw *SUVR* values overestimated small increases in Aβ load, and this bias increases at later time-windows. In comparison, the estimates based on *SUVR*c were time-stable and more accurate compared to *DVR*. Future work investigating amyloid reemergence after anti-amyloid therapies could benefit from improvements in quantifying small Aβ changes using *SUVR*c.

Methodologically, *SUVR* correction consists of estimating βtar followed by application of the nonlinear correction formula. A major contribution of this study is the development of empirical βtar models spanning the full spectrum of Aβ pathology. Building on previous work^19,20^, these models exploit the inverse relationship between regional radiotracer clearance and tracer uptake, while maintaining a simple implementation with radiotracer and time-window specific parameters that are shared across all regions, as a reasonable compromise between accuracy in βtar prediction against model simplicity and generalizability. As βtar estimation is independent of the correction formula itself, alternative approaches, such as neural networks trained on dynamic PET data, could also be explored.

A subtle issue pertains to the effect of frame-timing during the *SUVR* time-window on correction performance. In this work, we utilized 2-min frames with [^11^C]PiB data on ECAT HR+ while previous *SUVR*c work used 5-min frames on the HRRT with other radiotracers^19^. For an *SUVR* time-window of length T min with frame duration of Δt, image noise (s) is proportional to 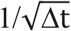 ^28^, and standard error of estimated parameters such as βcomp and βref is inversely proportional to 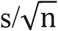 (n is number of time-points or n = T/ Δt). Therefore, standard error in estimated parameters is fairly independent of Δt, with higher noise due from a lower Δt balanced due to higher number of data points. We recommend future studies to use their usual OSEM settings and frame-durations (within a reasonable range of 1-5 min) when applying this method. Newer-generation high sensitivity PET systems with longer axial field-of-view can offer dramatic improvements in lowering image noise which may make direct estimation of βtar from TACs possible in future. A further consideration is the use of SRTM/SRTM2 *DVR* as the reference standard. Despite favorable test–retest variability^29^ and minimal bias relative to 2TCM *DVR* for [^11^C]PiB^9,30^, validation in larger cohorts with arterial sampling is recommended.

There are many tangible applications of this method to amyloid imaging that remain unexplored. An obvious application would be the extension of this method to other amyloid radiotracers such as [^18^F]Flutafuranol, [^18^F]Florbetapir, [^18^F]Flutemetamol and [^18^F]Florbetaben. Out of these, [^18^F]Flutafuranol kinetics are most similar to [^11^C]PiB^31^ and therefore, we expect the empirical models and estimated parameters developed here to generalize better to that tracer. *SUVR* correction can also be a step in harmonizing datasets across different amyloid tracers. Current methods for cross-tracer harmonization are based on converting *SUVR* values to the Centiloid scale, but correcting *SUVR* values prior to Centiloid conversion may help harmonize different time-window dependent non-equilibrium effects (kinetic harmonization).

In conclusion, we successfully adapted the *SUVR* correction approach to [^11^C]PiB imaging, demonstrating reductions in bias and variability of [^11^C]PiB *SUVR* with this method that can improve statistical power in cross-sectional studies and enable more accurate measurements of longitudinal amyloid change. We also present a roadmap of the key steps needed to implement this correction on future cohorts, covering the full spectrum of Aβ burden, making it possible to generalize this method to other cohorts.

## Data Availability

Datasets used here can be accessed from Harvard Aging Brain Study Public Dataset by submitting an application through the Synapse HABS Repository and agreeing to the Data Use Agreement (DUA).

