## Supplementary for "Translating *SUVR* bias correction to amyloid PET enables early imaging and more accurate simplified quantification"

### Supplementary Material

### Figures:

## A:

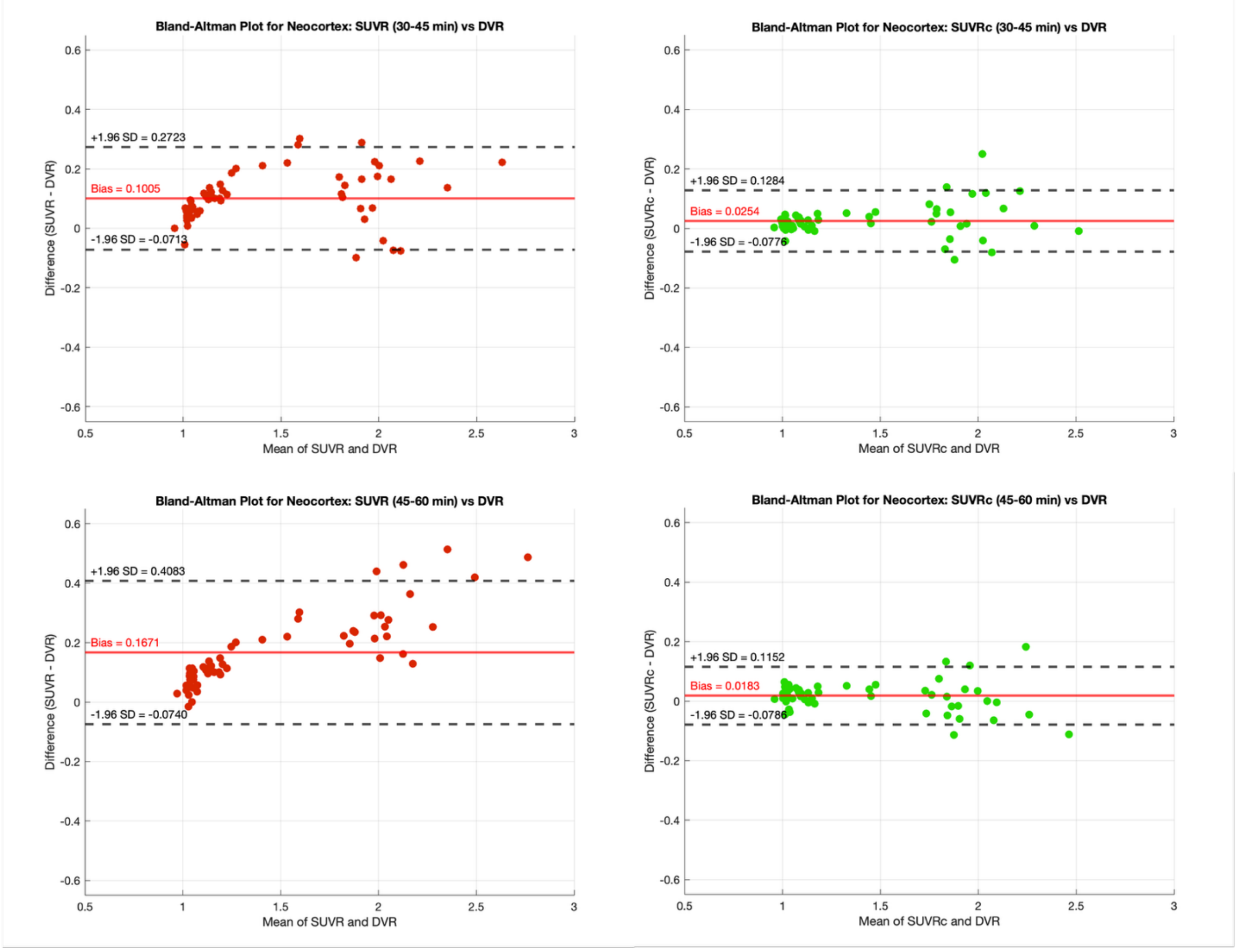

**B:**

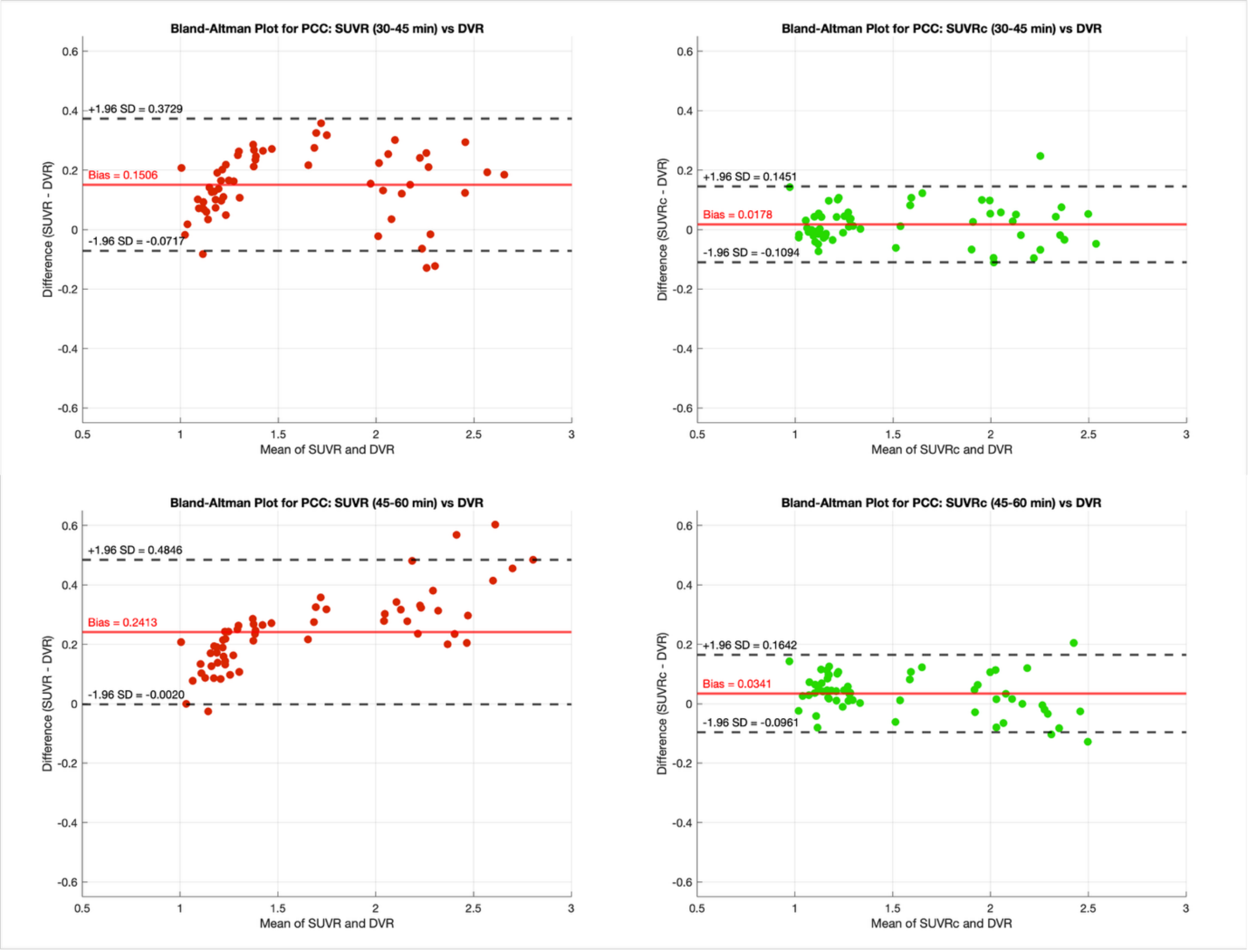


**Supplementary Figure 1:** Bland-Altman analyses at single-ROI level for the neocortex (global amyloid load, A) and posterior cingulate cortex (an early amyloid accumulation region, B) reveal similar trends seen in the main analysis pooling all regions together.

**A:**

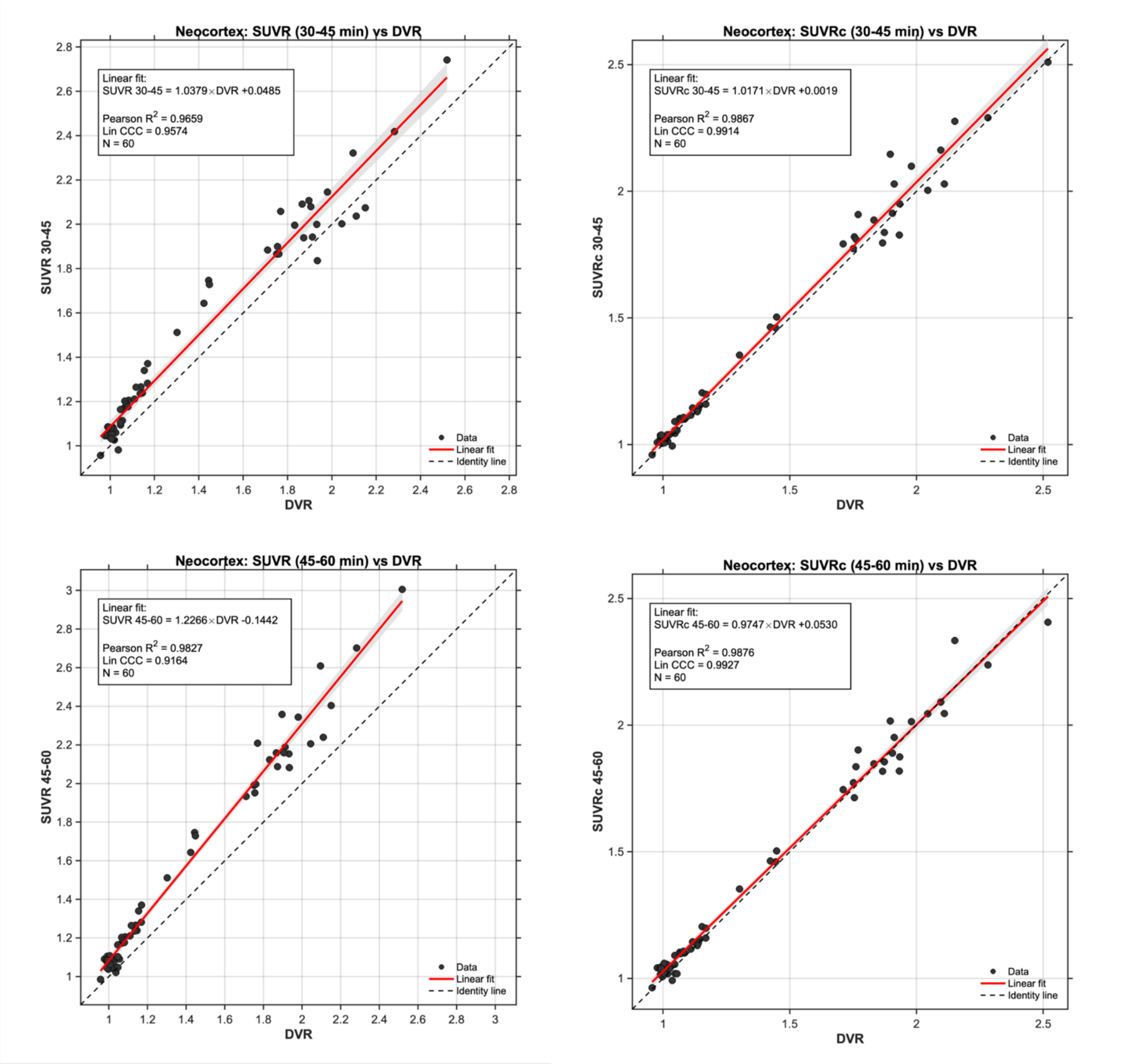
Neocortex: SUVR (30-45 min) vs DVR
Linear fit:
SUVR 30-45 = 1.0379×DVR +0.0485
Pearson $R^2$ = 0.9659
Lin CCC = 0.9574
N = 60
SUVR 30-45
DVR
Data
Linear fit
Identity line
Neocortex: SUVRc (30-45 min) vs DVR
Linear fit:
SUVRc 30-45 = 1.0171×DVR +0.0019
Pearson $R^2$ = 0.9867
Lin CCC = 0.9914
N = 60
SUVRc 30-45
DVR
Data
Linear fit
Identity line
Neocortex: SUVR (45-60 min) vs DVR
Linear fit:
SUVR 45-60 = 1.2266×DVR -0.1442
Pearson $R^2$ = 0.9827
Lin CCC = 0.9164
N = 60
SUVR 45-60
DVR
Data
Linear fit
Identity line
Neocortex: SUVRc (45-60 min) vs DVR
Linear fit:
SUVRc 45-60 = 0.9747×DVR +0.0530
Pearson $R^2$ = 0.9876
Lin CCC = 0.9927
N = 60
SUVRc 45-60
DVR
Data
Linear fit
Identity line

**B:**

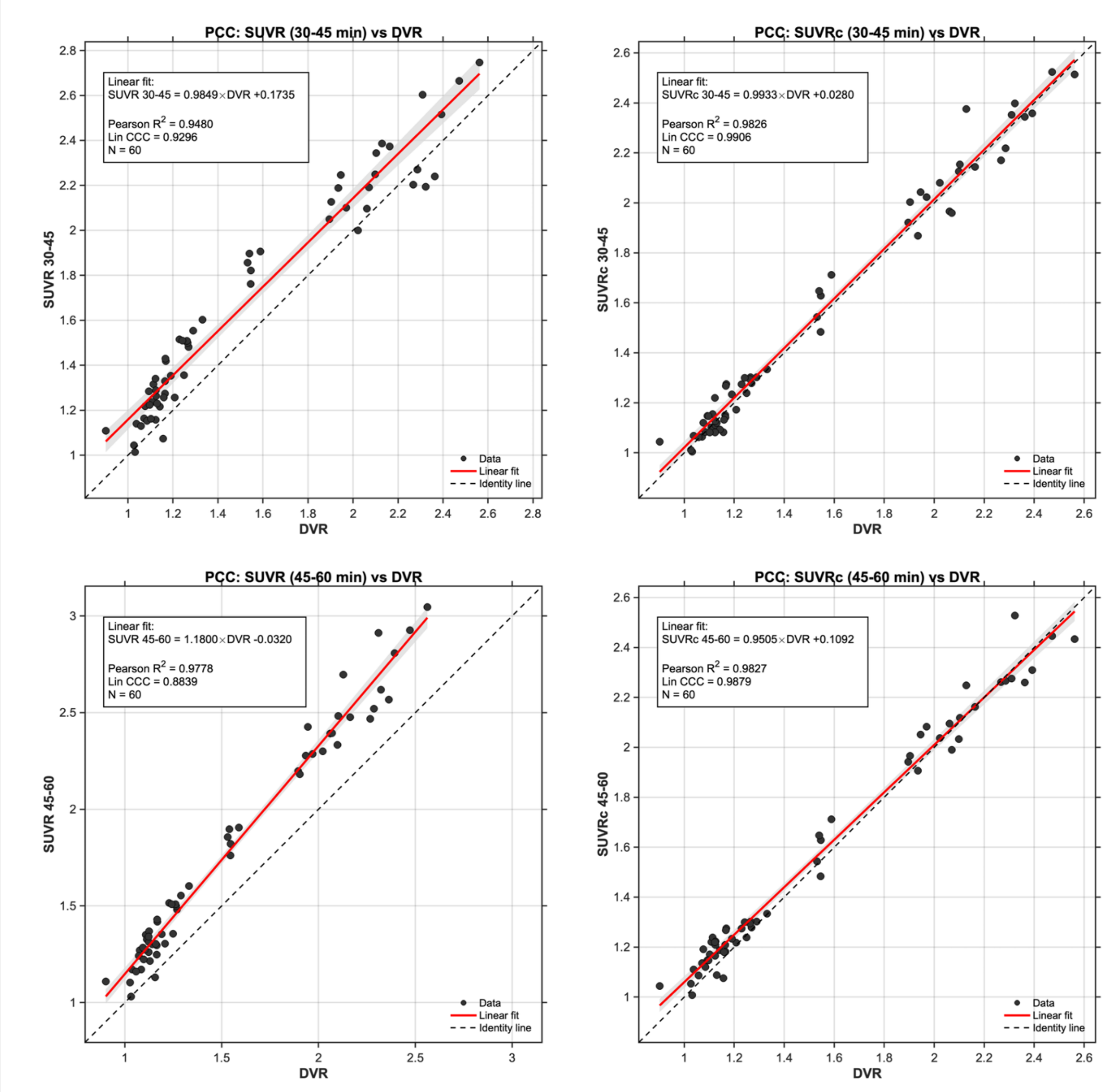


**Supplementary Figure 2:** Pearson's correlation coefficient and Lin's concordance coefficient (LCC) analyses at single-ROI level for the neocortex (global amyloid load, A) and posterior cingulate cortex (an early amyloid accumulation region, B) reveal similar trends seen in the main analysis pooling all regions.

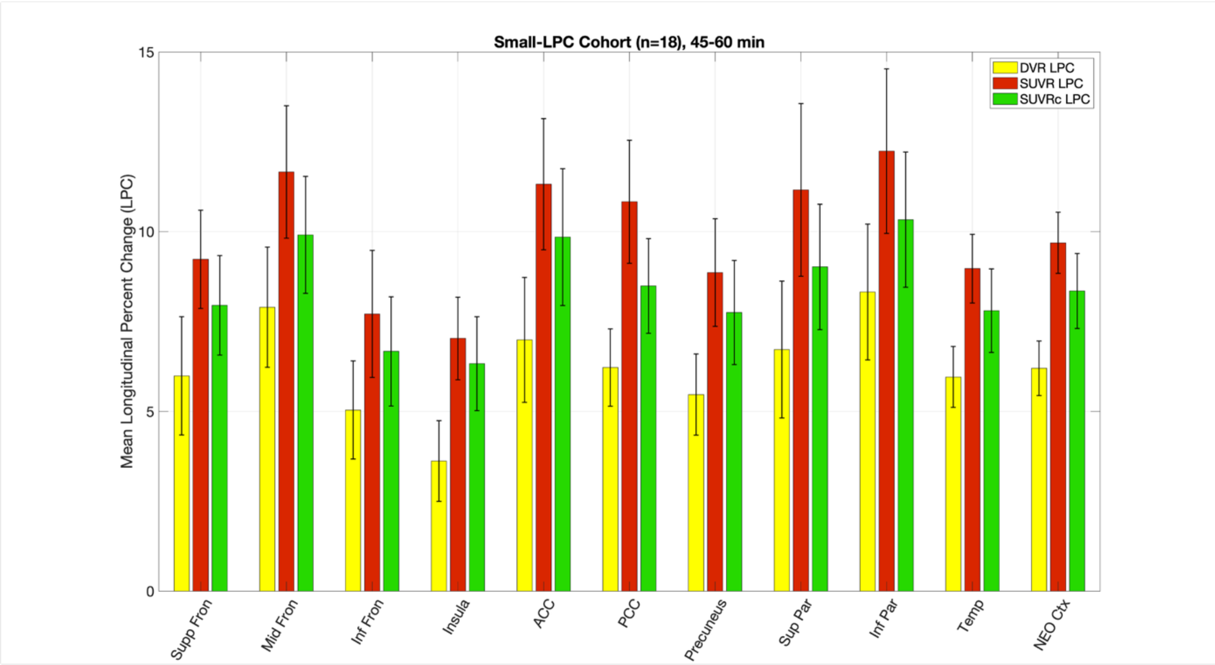


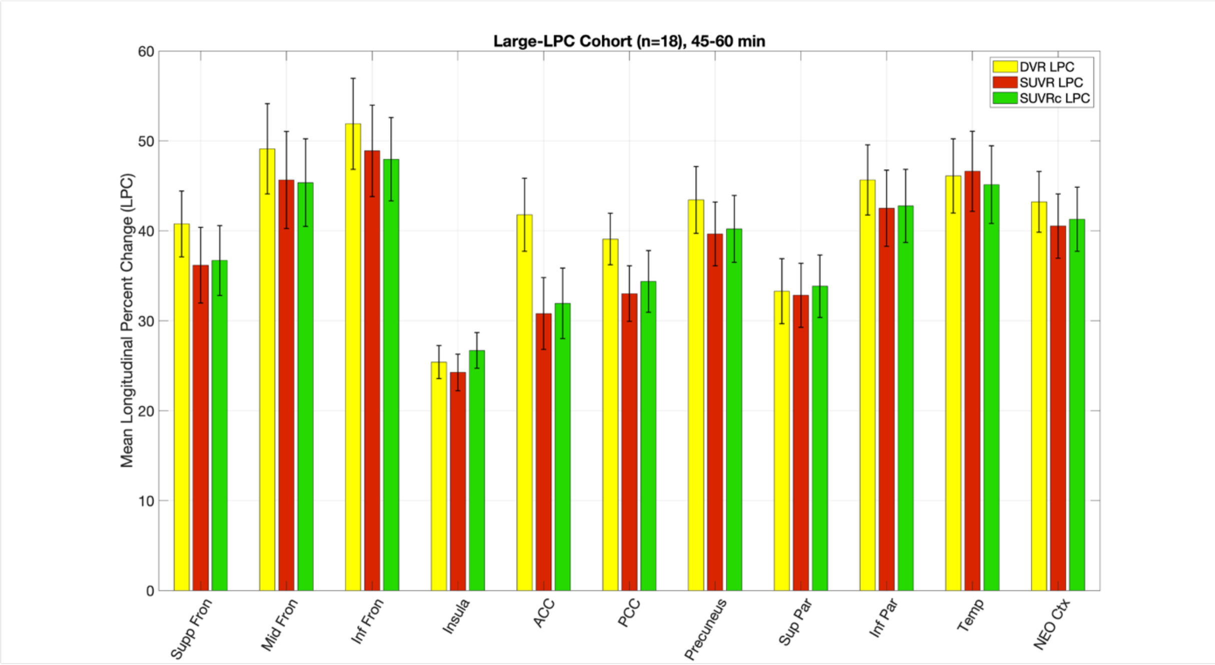


**Supplementary Figure 3:** Mean longitudinal percent change values (mean ± s.e.m) for *SUVR* and *SUVR*c at 45-60 min show that both metrics perform similarly for large longitudinal changes, but *SUVR* tends to overestimate smaller longitudinal changes more at this time-window. *SUVR*c also overestimates small longitudinal changes substantially though not as much as *SUVR*. For longitudinal imaging, as well as for general participant comfort, we recommend 30-45 min *SUVR*c as the optimal metric since it closely follows changes in *DVR*.

### Implementation Details for Simplified Reference Tissue Model Fits to PET Data

For each individual, TACs (0-60 min) from target and reference regions were fit to the SRTM in a single step to estimate non-displaceable binding potentials ($BP_{ND}$) and relative tracer delivery ($R_1$) values for the target regions, along with a single estimate for $k_2$' of the reference region. For scans with low and intermediate amyloid-burden, SRTM fits often fail due to degeneracy in estimated values of $BP_{ND}$ and $k_2$' (several values of $k_2/k_2$' ratio can simultaneously describe a low amyloid time-activity curve [TAC]). Therefore, for these scans $k_2$' of the cerebellar cortex was first estimated by fitting the TAC of eroded cerebral white matter with SRTM. The higher retention of [$^{11}$C]PiB in cerebral white matter leads to a distinct TAC compared to the reference region, allowing a robust estimate of subject-level $k_2$' value as has been previously reported[1]. This fixed $k_2$' value was then used in SRTM2[2] to estimate $BP_{ND}$ and $R_1$ values for target regions. In the absence of arterial blood data, ground-truth regional *DVR* values for all scans were computed as $BP_{ND}$ + 1 from SRTM or SRTM2 fits, as described.

As an example, consider participant #18 from the low amyloid cross-sectional cohort. This participant had very low levels of brain amyloid and therefore, TACs from all regions were very similar to the reference region. Supplementary Figure 4 shows the TACs for the reference region (cerebellar gray) and PCC (region with most amyloid) for this participant:

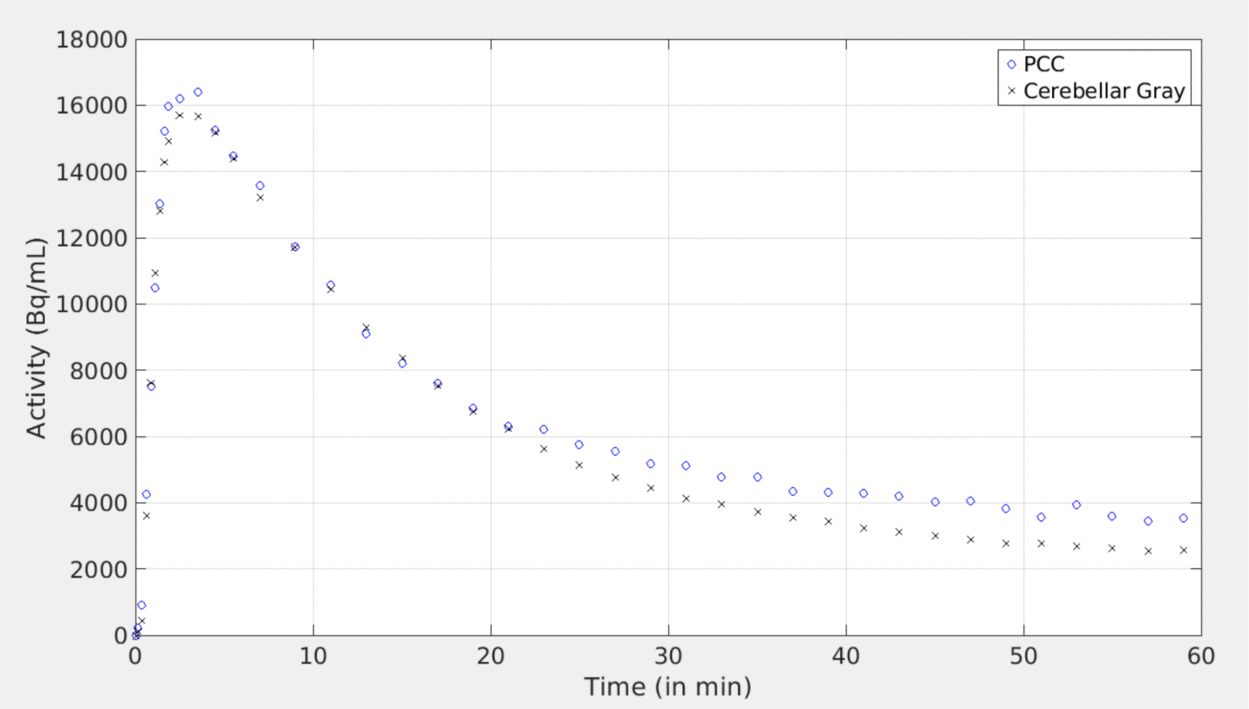


**Supplementary Figure 4:** TACs of reference and a target region with most amyloid (posterior cingulate) for a participant with overall low amyloid levels.

For this participant, the basic SRTM fit resulted in the following values of estimated parameters ± se: $k_2$': 0.0026 ± 0.005 min$^{-1}$, $BP_{ND}$: 27.8 ± 1368, $R_1$: 1.01 ± 0.006. $k_2$' is severely underestimated here, along with a corresponding high yet incorrect estimate for $BP_{ND}$, even though the SRTM fit to the TAC looks acceptable (Supplementary Figure 5).

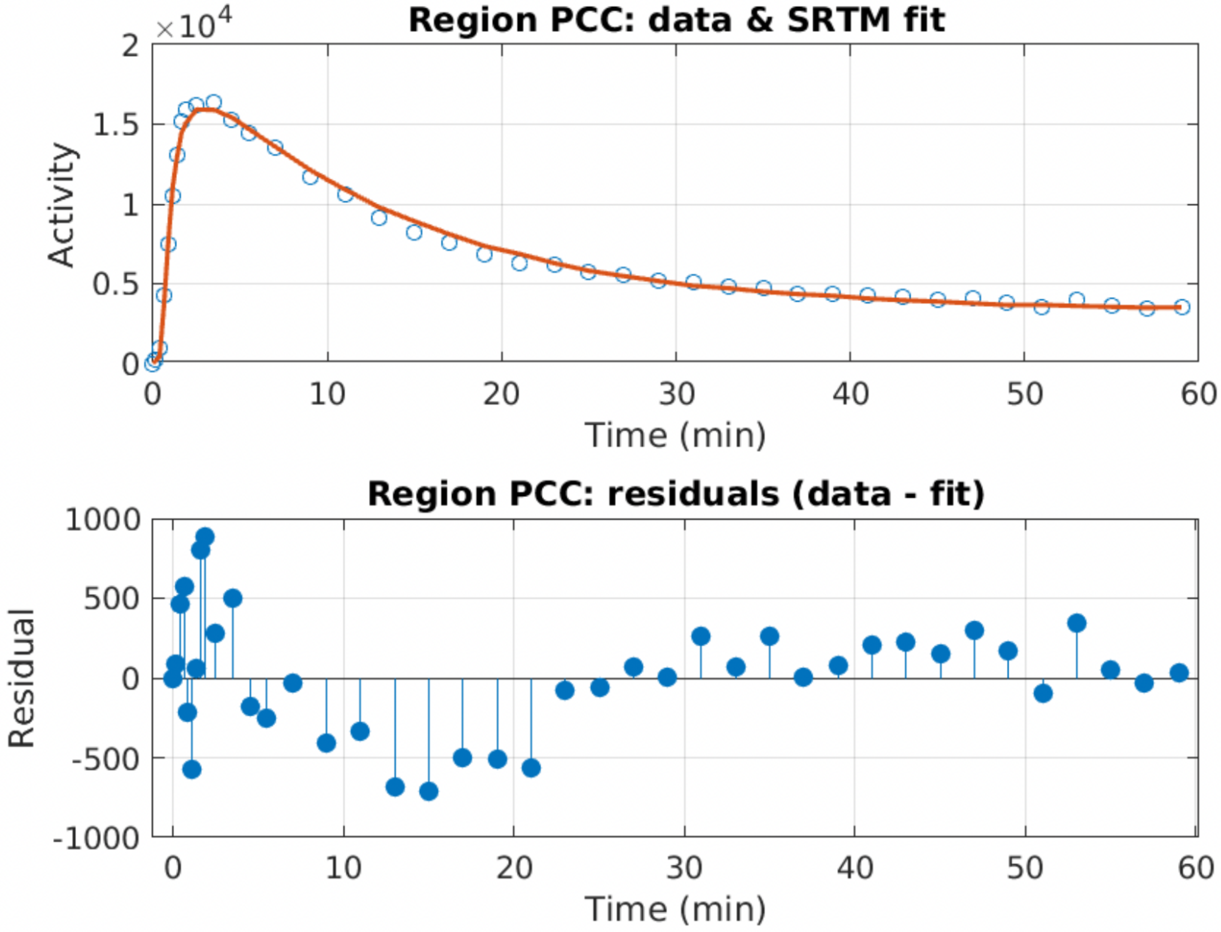


**Supplementary Figure 5:** SRTM fit to the PCC of the low-amyloid participant is visually acceptable despite degeneracy issues leading to incorrect values of $k_2$' and $BP_{ND}$.

In this case, it is better to first estimate $k_2$' of cerebellar gray matter (reference region) by first fitting SRTM to the eroded cerebral white-matter region which has a distinctly different TAC shape and magnitude than that of the reference region (Supplementary Figure 6). This led to an estimated value of $k_2$' of 0.052 ± 0.001 min$^{-1}$, a value more aligned with known kinetics of [$^{11}$C]PiB and ~ a 20-fold increase from the previously estimated low value of 0.0026 min$^{-1}$.

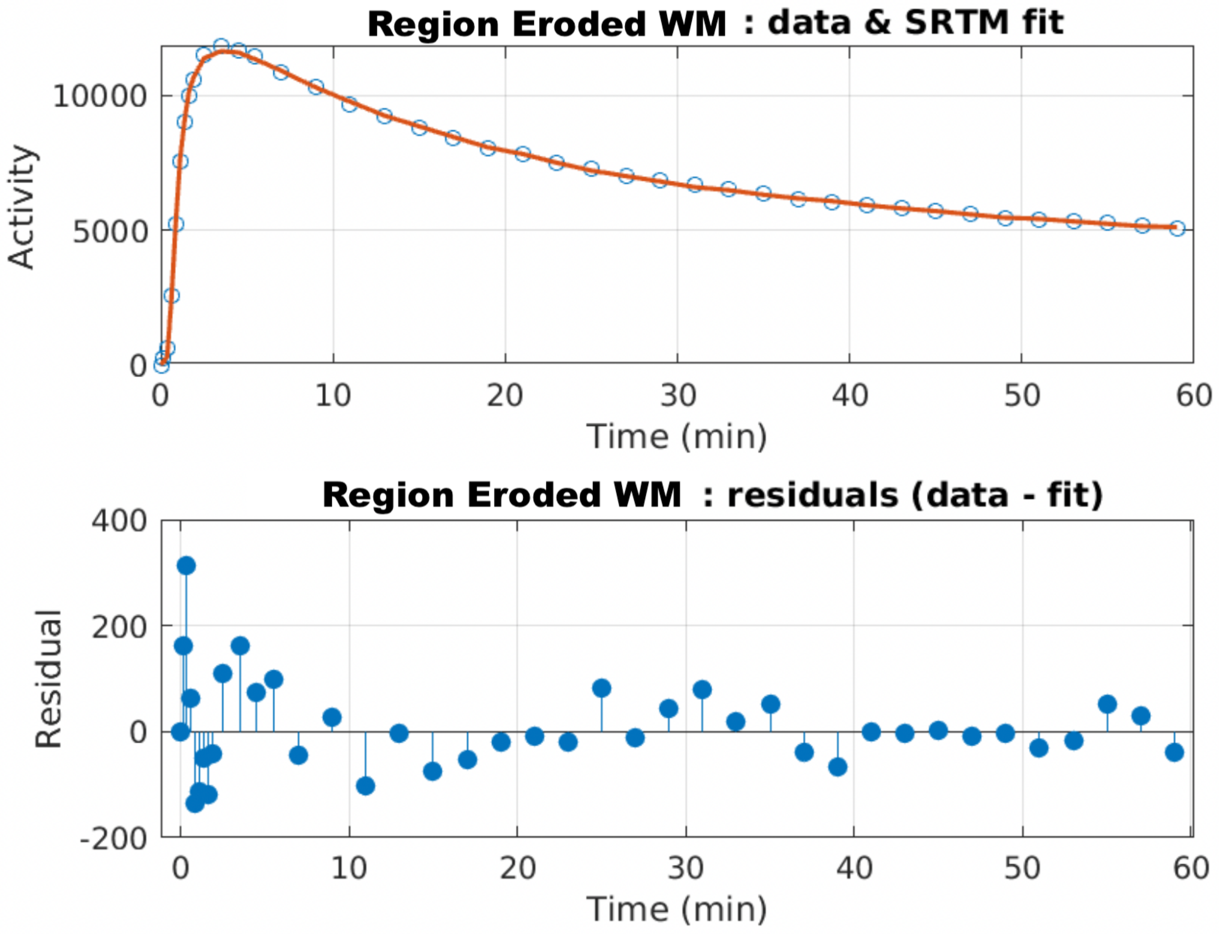


**Supplementary Figure 6:** SRTM fit to the eroded cerebral white-matter of the low-amyloid participant. While WM is not a target region-of-interest it can be used to estimate a more accurate value of $k_2$' in low-amyloid participants.

With the fixed value of $k_2$' = 0.052 $min^{-1}$, SRTM2 can be used to fit all target TACs and estimate $BP_{ND}$ and $R_1$ values. For example, the fit in the PCC now yields more realistic values for $BP_{ND}$: 0.13 ± 0.01 and $R_1$: 1.00 ± 0.007. The final SRTM2 fit is shown in Supplementary Figure 7.

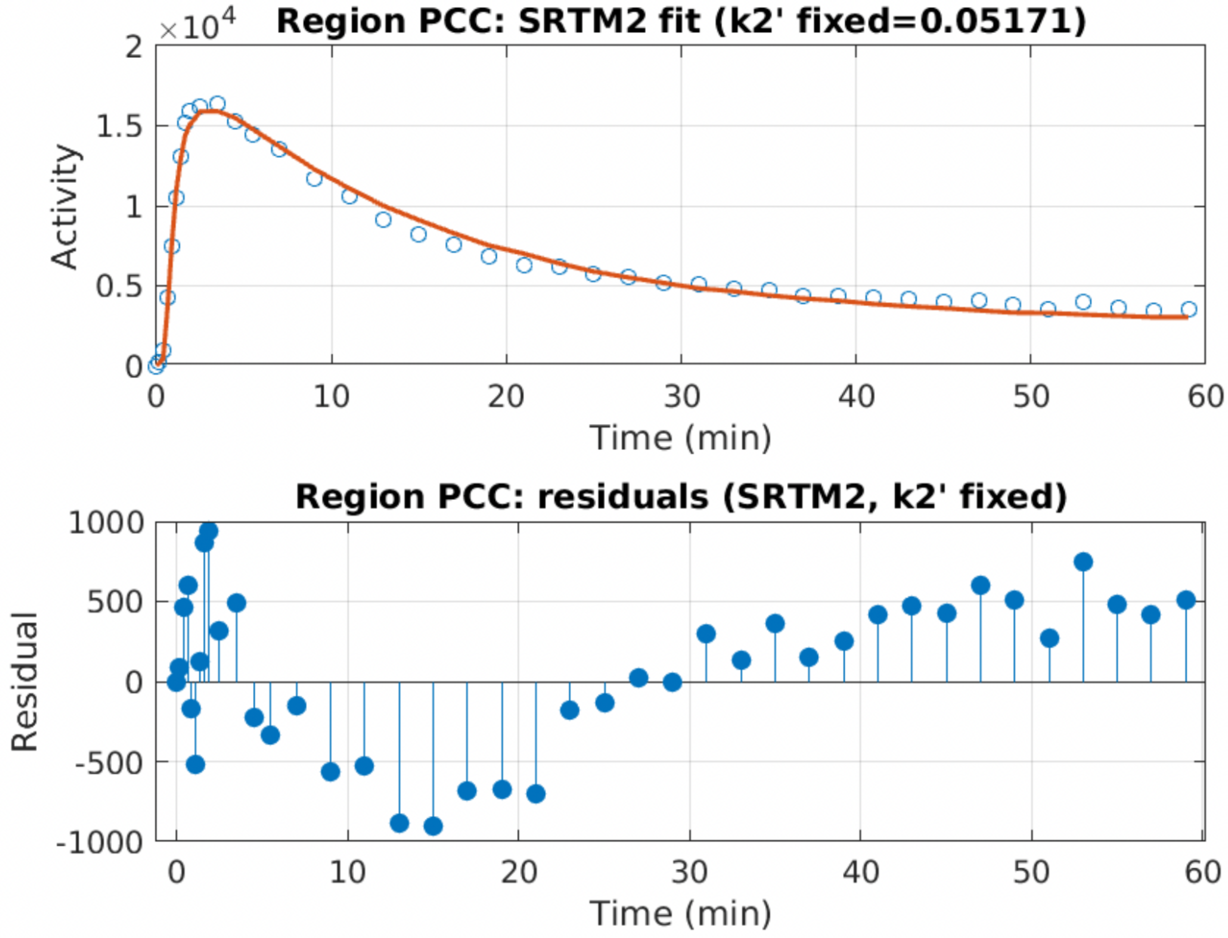


**Supplementary Figure 7:** SRTM2 fit to PCC TAC for the low amyloid participant using a fixed value of $k_2$' = 0.052 min$^{-1}$ that was estimated by fitting cerebral white-matter TAC.

Finally, Supplementary Figure 8 shows the estimated values of $k_2$' by fitting SRTM to eroded cerebral white-matter TACs across all participants (training + validation) in the three cross-sectional cohorts. Note that the $k_2$' values were not significantly different between any groups as expected from derived metrics of the reference region.

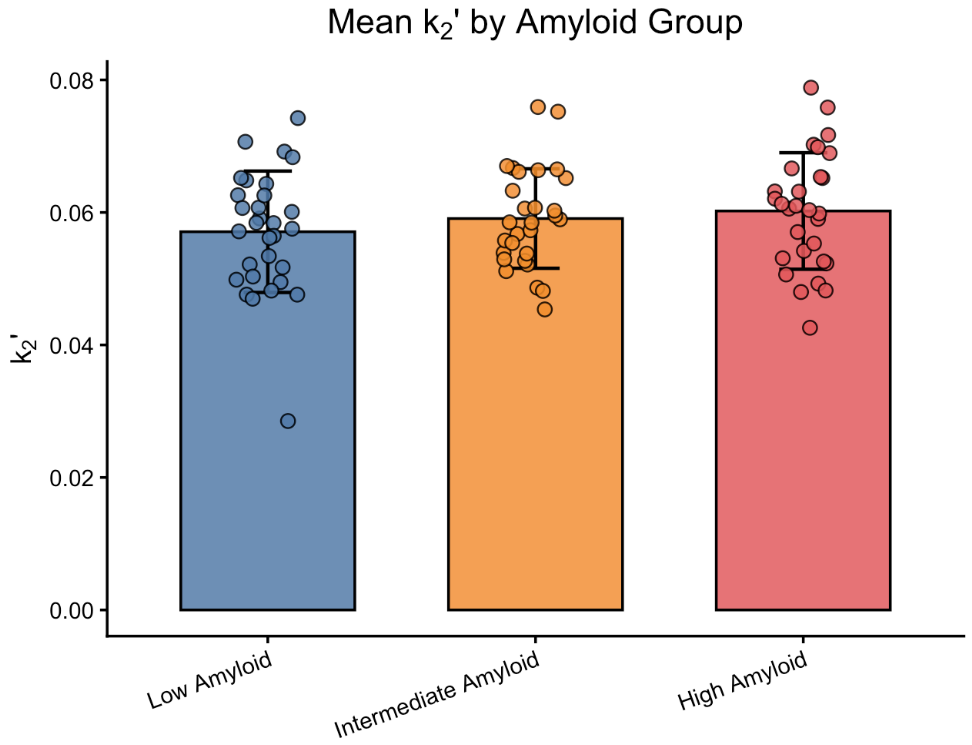


**Supplementary Figure 8:** Estimated values of $k_2$' for the cerebellar gray matter (reference region) in the low, intermediate and high amyloid groups using SRTM fits to eroded cerebral white-matter TACs. None of the groups showed significant differences (2-tailed unpaired *t*-tests). Note that we did not use these estimates of $k_2$' for the high amyloid group as direct SRTM fits generated reliable $k_2$' and $BP_{ND}$ values in this cohort but they are nonetheless presented here for comparison.


1 Peretti, D. E. R., F.E.; Doorduin, J.; De Jong, B.M.; De Deyn; P.P., Dierckx; R.A., Boellaard, R.; Vállez García, D. Optimization of the k 2' parameter estimation for the pharmacokinetic modeling of dynamic PIB PET scans using SRTM2. *Frontiers in Physics* **7**, 212 (2019).

2 Wu, Y. & Carson, R. E. Noise reduction in the simplified reference tissue model for neuroreceptor functional imaging. *J Cereb Blood Flow Metab* **22**, 1440–1452 (2002). https://doi.org/10.1097/01.WCB.0000033967.83623.34
